# What do women know about breast density? A public screening program perspective

**DOI:** 10.1101/2020.04.02.20048371

**Authors:** Monique Robertson, Ellie C Darcey, Evenda K Dench, Louise Keogh, Kirsty McLean, Sarah Pirikahu, Christobel Saunders, Sandra Thompson, Catherine Woulfe, Elizabeth Wylie, Jennifer Stone

## Abstract

**Background:** This study assesses knowledge of breast density, one of breast cancer’s strongest risk factors, in women attending a public mammographic screening program in Western Australia that routinely notifies women if they have dense breasts.

**Methods:** Survey data was collected from women who were notified they have dense breasts and women who had not (controls). Descriptive data analysis was used to summarize responses.

**Results:** Of the 6183 women surveyed, over 85% of notified women knew that breast density makes it difficult to see cancer on a mammogram (53.9% in controls). A quarter of notified women knew that having dense breasts puts women at increased risk for breast cancer (13.2% in controls). Overall, 50.1% of notified women indicated that they thought the amount of information provided was “just right” and 24.9% thought it was “too little”, particularly women notified for the first time (32.1%).

**Conclusion:** The main message of reduced sensitivity of mammography in women with dense breasts provided by the screening program appears to be getting though. However, women are largely unaware that increased breast density is associated with increased risk. Women notified of having dense breasts for the first time could potentially benefit from additional information.

## Introduction

Researchers have been investigating the associations between breast density and breast cancer for many years, but the concept of breast density is still relatively new to consumers of breast screening services. It is unclear whether many women know that breast density is a strong predictor of breast cancer risk (1) or that it significantly reduces the sensitivity of mammography, making the detection of tumours more challenging at the time of screening (2). It is also unknown whether women understand that breast density cannot be determined by size, feel or touch - it is typically measured via mammography and it is estimated that around 43% of screening populations aged 40-74 years having heterogeneously or extremely dense breasts (Breast Imaging-Reporting and Data System (BIRADS) categories c or d) (3). Although dense breast tissue is common, only the minority of women with dense breasts will develop breast cancer. Identifying those who are at higher risk is challenging and the increasing benefits of supplemental screening is accompanied by an increase in harms and costs. The Society of Breast Imaging appropriateness criteria recommends that women at intermediate risk of breast cancer due to a family history, a personal history of breast cancer, or other risk factors may benefit from regular supplemental breast ultrasound (4). Lack of evidence around the most effective screening recommendations for women with dense breasts makes informing women about what to do about breast density difficult.

Legislation in the United States (US) mandating that all screened women be informed of their breast density has increased consumer advocacy and pressure for other screening programs internationally to routinely measure and report breast density to participants. A recent review of the impact of breast density notification legislation highlighted that current evidence is predominantly from retrospective single institution studies (comparing pre- and post-legislation) or small surveys, with very few population-level studies (5). In early 2019, the US Congress directed the Food and Drug Administration (FDA) to establish a national minimum standard for breast density notification as part of routine mammography reporting. Therefore, population-based evidence of what women know about breast density and how well women interpret breast density information provided by screening services is needed to inform and improve breast density notification standards.

BreastScreen Western Australia (WA) is a public breast cancer screening program that has been notifying women if they have dense breasts for over a decade. Density measurement is dichotomous; a radiologist assessment of a BIRADS category 3 or 4 (4th edition) is considered dense. For women flagged as having dense breasts, the routine results letter states that mammograms are significantly less sensitive in diagnosing breast cancer in women with dense breasts and recommends they consult their General Practitioner (GP). Sample text is provided in Supplementary Figure 1. BreastScreen WA also provides the results letter to their nominated GP and links for additional information via BreastScreen WA’s website.

This study is the first to directly assess the knowledge of breast density between women who were notified of having dense breasts and controls (who were not flagged as having dense breasts) within the BreastScreen WA program via a survey of nearly 7000 participants who received a routine results letter after their most recent screening mammogram. Notified women were also assessed on how well they understood the information provided.

## Methods

### Survey development

A consumer consultation organized by WA’s Consumer Community Health Research Network was held to gain insight into key themes to be included in the survey. A comprehensive literature review generated an extensive list of validated questions to address each of the themes. Questions were reviewed and shortlisted by the BreastScreen WA Consumer Reference Group and the Cancer Epidemiology Network’s Consumer Advisory Council. This article presents an analysis of women’s responses to the 12 survey questions designed to elicit data on women’s knowledge of breast density and reactions to receiving dense breast notification. The software program Qualtrics was used to create an online version. The survey officially launched via email invitation on December 19th, 2017.

Ethics and governance approval was obtained from King Edward Memorial Hospital’s Human Research Ethics Committee (#2017046EW RGS000474) and The University of Western Australia RA/4/20/4178).

### Recruitment

All women who attended BreastScreen WA from 21 November 2017 to 19 April 2018 and who received a routine results letter were emailed an invitation to participate in the survey. Breast density notification is included as additional information within the routine results letter for women flagged as having dense breasts by at least one radiologist; otherwise breast density information is not provided. Women who did not provide an email address were not invited to participate (∼30%). Women had the option of completing the survey either online or by telephone. The online survey took approximately 5-10 minutes. Breast density notification status was known but responses were otherwise anonymous unless women opted to provide their contact details for future contact.

### Selection of subjects

Respondents were categorised into three groups based on their most recent results letter *and* their response to the question “Have you ever been told by a health professional (e.g. BreastScreen WA) that your breast tissue is dense?”: controls, notified for the first time, notified more than once. See Supplemental Figure 2. Controls were defined as women who were not flagged/notified as having dense breasts and who responded either “No” or “I don’t know” (n=3008). The “notified” groups were defined as women who were notified as having dense breasts *and* who responded that this was the first time they were told (n=1381) or that they had been told more than once (n=1308). Henceforth “notified” women refers collectively to these groups. To ensure responses truly reflect the defined groups, women who did not fall into these three groups were excluded from the analysis (n=1421). Women whose responses were excluded were more likely to be Caucasian/European and have a slightly higher socio-economic status (data not shown).

Demographic information collected included year of birth, ethnicity and postcode to determine the Australian Bureau of Statistics’ Accessibility/Remoteness Index for Areas (ARIA) and Socio-Economic Indexes for Areas (SEIFA) (6). Respondents whose age was younger than the BreastScreen WA screening age (40 years) were also excluded (n=4).

### Statistical methods

All data management and quantitative analyses were undertaken using Stata v14. Descriptive statistics were used to describe the survey responses of the three groups (control group, notified for the first time, notified multiple times). Logistic regression was used to examine predictors of knowledge, particularly knowledge of the reduced sensitivity of mammography (“masking”) in notified women. Knowledge of masking was defined as “Yes” responses to the question “Do you think dense breast tissue makes it more difficult to see cancer on a mammogram?” compared to “Unsure” or “No” responses. Covariates investigated as potential predictors of knowledge of masking included age, ethnicity, remoteness of residence, education and occupation, and number of times notified. See Supplemental Table 2.

### Thematic analysis

Women were provided an opportunity to comment on the information learned via an open-ended question. Responses were analysed thematically through an inductive approach to summarise the full range of responses into a meaningful framework, and counts were used to gain a sense of both the spread and patterns in responses (7).

## Results

Overall, 30 566 women were emailed the survey invitation, 1150 emails bounced back and 72 women unsubscribed. Of the 29 416 who received email invitations, 6922 responded to the survey resulting in a 23.5% response rate (See Supplemental Figure 2). The response rate was higher within women who were notified they have dense breasts (28.1%) compared to those who were not notified (19.8%).

Characteristics of respondents by defined groups are shown in Supplemental Table 1. Most respondents were aged between 50 and 74 years, were Caucasian/European, resided in major cities and were generally in higher quintiles of the socio-economic indices. Women who had been notified for the first time were younger than those who had been notified multiple times and the controls.

**Table 1.** Frequency of responses regarding respondents’ awareness of breast density for control and notified women.

| Survey questions | Controls<br>(n=3008) | Notified for<br>first time<br>(n=1381) | Notified<br>multiple times<br>(n=1308) |
| --- | --- | --- | --- |
| Have you ever heard of breast density? /Prior to being told that your breast tissue is dense had you ever heard of breast density?<sup>a</sup> (%) |  |  |  |
| No | 1027 (34.1%) | 697 (50.5%) | 646 (49.4%) |
| Yes | 1844 (61.3%) | 643 (46.6%) | 625 (47.8%) |
| I don't know | 132 (4.4%) | 35 (2.5%) | 37 (2.8%) |
| Missing | 5 (0.2%) | 6 (0.4%) | 0 (0.0%) |
| Tell us what you understand your own breast density to be? (%) | 1844<sup>b</sup> | 1375<sup>c</sup> | 1308<sup>c</sup> |
| Lower than average | 114 (6.2%) | 35 (2.6%) | 19 (1.5%) |
| About average | 697 (37.8%) | 164 (11.9%) | 212 (16.2%) |
| Higher than average | 65 (3.5%) | 644 (46.8%) | 758 (58.0%) |
| I have no idea | 965 (52.3%) | 532 (38.7%) | 319 (24.4%) |
| Missing | 3 (0.16%) | 0 (0.0%) | 0 (0.0%) |
| Where did this information/advice come from?<sup>e</sup> (%) | 876<sup>d</sup> | n/a | n/a |
| Self-assumption | 289 (33.0) |  |  |
| Breast screening services | 201 (23.0) |  |  |
| Health service providers | 138 (15.8) |  |  |
| Outside health service | 142 (16.2) |  |  |
| Unsure | 37 (4.2) |  |  |
| Other | 15 (1.7) |  |  |
| Missing | 71 (8.1) |  |  |
| If you are not aware of your breast density, would you like to know your breast density? (%) | 965<sup>f</sup> | n/a | n/a |
| Yes | 699 (72.4%) |  |  |
| No | 58 (6.0%) |  |  |
| I don't know | 182 (18.9%) |  |  |
| Missing | 26 (2.7%) |  |  |

<sup>a</sup>Survey question differed between controls ('Have you ever heard of breast density?') and notified women ('Prior to being told that your breast tissue is dense had you ever heard of breast density?'); <sup>b</sup>For controls, the question was only asked for those women who answered 'Yes' to 'Have you ever heard of breast density?'; <sup>c</sup>For notified women, the question was not asked of those who did not answer the question 'Prior to being told that your breast tissue is dense had you ever heard of breast density?'; <sup>d</sup>Only controls who answered 'Lower than average', 'About average' or 'higher than average' to the question 'Tell us what you understand your own breast density to be?' were asked this question; <sup>e</sup>Responses could be given more than one code; <sup>f</sup>Only controls who answered 'I have no idea' to the question 'Tell us what you understand your own breast density to be?' were asked this question. (**abbreviations:** n/a= not applicable)

**Table 2.**
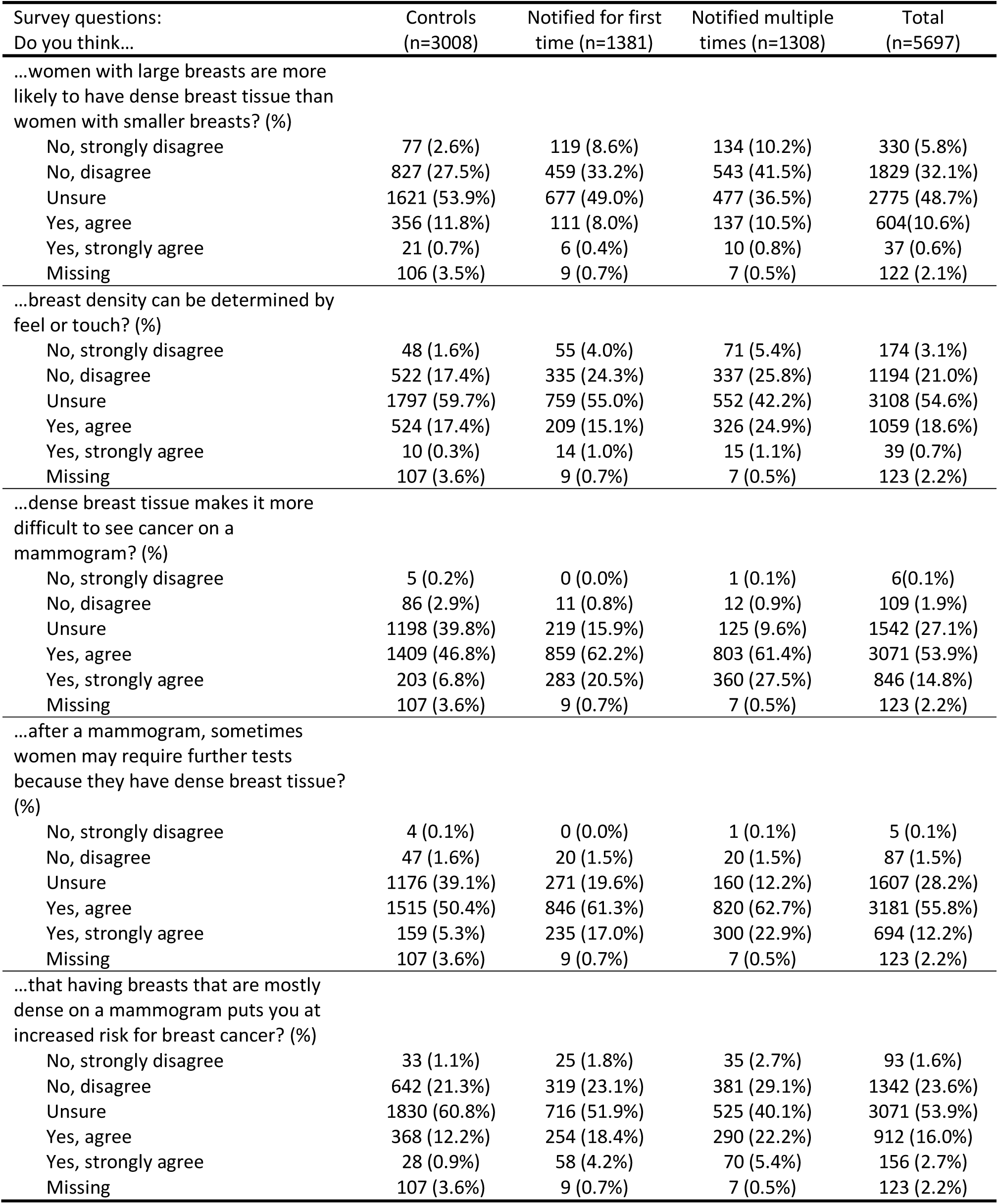
Frequency of responses regarding general knowledge of breast density for control and notified women.

| Survey questions:<br>Do you think... | Controls<br>(n=3008) | Notified for first<br>time (n=1381) | Notified multiple<br>times (n=1308) | Total<br>(n=5697) |
| --- | --- | --- | --- | --- |
| ...women with large breasts are more likely to have dense breast tissue than women with smaller breasts? (%) |  |  |  |  |
| No, strongly disagree | 77 (2.6%) | 119 (8.6%) | 134 (10.2%) | 330 (5.8%) |
| No, disagree | 827 (27.5%) | 459 (33.2%) | 543 (41.5%) | 1829 (32.1%) |
| Unsure | 1621 (53.9%) | 677 (49.0%) | 477 (36.5%) | 2775 (48.7%) |
| Yes, agree | 356 (11.8%) | 111 (8.0%) | 137 (10.5%) | 604 (10.6%) |
| Yes, strongly agree | 21 (0.7%) | 6 (0.4%) | 10 (0.8%) | 37 (0.6%) |
| Missing | 106 (3.5%) | 9 (0.7%) | 7 (0.5%) | 122 (2.1%) |
| ...breast density can be determined by feel or touch? (%) |  |  |  |  |
| No, strongly disagree | 48 (1.6%) | 55 (4.0%) | 71 (5.4%) | 174 (3.1%) |
| No, disagree | 522 (17.4%) | 335 (24.3%) | 337 (25.8%) | 1194 (21.0%) |
| Unsure | 1797 (59.7%) | 759 (55.0%) | 552 (42.2%) | 3108 (54.6%) |
| Yes, agree | 524 (17.4%) | 209 (15.1%) | 326 (24.9%) | 1059 (18.6%) |
| Yes, strongly agree | 10 (0.3%) | 14 (1.0%) | 15 (1.1%) | 39 (0.7%) |
| Missing | 107 (3.6%) | 9 (0.7%) | 7 (0.5%) | 123 (2.2%) |
| ...dense breast tissue makes it more difficult to see cancer on a mammogram? (%) |  |  |  |  |
| No, strongly disagree | 5 (0.2%) | 0 (0.0%) | 1 (0.1%) | 6 (0.1%) |
| No, disagree | 86 (2.9%) | 11 (0.8%) | 12 (0.9%) | 109 (1.9%) |
| Unsure | 1198 (39.8%) | 219 (15.9%) | 125 (9.6%) | 1542 (27.1%) |
| Yes, agree | 1409 (46.8%) | 859 (62.2%) | 803 (61.4%) | 3071 (53.9%) |
| Yes, strongly agree | 203 (6.8%) | 283 (20.5%) | 360 (27.5%) | 846 (14.8%) |
| Missing | 107 (3.6%) | 9 (0.7%) | 7 (0.5%) | 123 (2.2%) |
| ...after a mammogram, sometimes women may require further tests because they have dense breast tissue? (%) |  |  |  |  |
| No, strongly disagree | 4 (0.1%) | 0 (0.0%) | 1 (0.1%) | 5 (0.1%) |
| No, disagree | 47 (1.6%) | 20 (1.5%) | 20 (1.5%) | 87 (1.5%) |
| Unsure | 1176 (39.1%) | 271 (19.6%) | 160 (12.2%) | 1607 (28.2%) |
| Yes, agree | 1515 (50.4%) | 846 (61.3%) | 820 (62.7%) | 3181 (55.8%) |
| Yes, strongly agree | 159 (5.3%) | 235 (17.0%) | 300 (22.9%) | 694 (12.2%) |
| Missing | 107 (3.6%) | 9 (0.7%) | 7 (0.5%) | 123 (2.2%) |
| ...that having breasts that are mostly dense on a mammogram puts you at increased risk for breast cancer? (%) |  |  |  |  |
| No, strongly disagree | 33 (1.1%) | 25 (1.8%) | 35 (2.7%) | 93 (1.6%) |
| No, disagree | 642 (21.3%) | 319 (23.1%) | 381 (29.1%) | 1342 (23.6%) |
| Unsure | 1830 (60.8%) | 716 (51.9%) | 525 (40.1%) | 3071 (53.9%) |
| Yes, agree | 368 (12.2%) | 254 (18.4%) | 290 (22.2%) | 912 (16.0%) |
| Yes, strongly agree | 28 (0.9%) | 58 (4.2%) | 70 (5.4%) | 156 (2.7%) |
| Missing | 107 (3.6%) | 9 (0.7%) | 7 (0.5%) | 123 (2.2%) |

Table 1 summarizes women’s responses regarding awareness of breast density. Over 60% of controls had heard of breast density. The majority of controls indicated they had “no idea” of their breast density (52.3%) or understood their density to be about average (37.8%), whereas notified women most commonly responded that they understood their density to be higher than average (52.1%). Among controls, the most commonly reported sources of the density information were self-assumption (33%) (e.g., “a guess”, “my own assumption”). When controls who were not aware of their breast density were asked if they would like to know their breast density, 72.4% indicated yes.

Table 2 summarises responses to the five knowledge questions. Only 19% of respondents overall agreed that breast density increased breast cancer risk, controls having the lowest agreement (13%). Overall, ‘unsure’ was the most commonly reported response to questions about whether density is related to breast size (49%) or breast cancer risk (54%), and whether it can be determined by touch (55%). A majority of respondents agreed that density decreases mammogram sensitivity (69%) and that women with dense breasts require further tests after a mammogram (68%). Agreement with these two statements were lowest in controls and highest in women notified multiple times.

The results of the logistic regression examining predictors of knowledge of masking show that older women were less likely to be knowledgeable (OR= 0.985, 95%CI=0.972, 0.998). Compared to women identifying as Caucasian/European, women who identified themselves as Asian ethnicity were less likely to be knowledgeable regarding masking (OR=0.264, 95%CI=0.172, 0.406). Women residing in remote or very remote areas and those in lower quintiles of education and occupation, were less likely to have knowledge of masking compared to the respective reference populations. Knowledge of masking was more likely among women who had been notified multiple times compared to those who had been notified for the first time (OR=1.82, 95%CI=1.43, 2.31) (see Supplementary Table 2).

Women’s responses regarding the information provided by BreastScreen WA (Table 3) indicated that over 60% had read the information provided by BreastScreen WA completely whilst 11% reported that they did not read it at all. Also, from Supplemental Figure 2, 14% of women whose results letter included breast density notification indicated either that they had not been told or did not know if they had been told they have dense breasts, suggesting that a substantial proportion of women don’t receive, read or comprehend the results letter. Overall, 52% of notified women thought the information provided by BreastScreen WA was “just right” whilst 25% of women thought it was “too little”, particularly women who were notified for the first time (32%).

**Table 3.** Frequency of responses regarding the information provided by BreastScreen WA for notified women.

| Survey questions | Notified for first time<br>(n=1381) | Notified multiple times<br>(n=1308) | Total<br>(2689) |
| --- | --- | --- | --- |
| The first time BreastScreen WA told you that your breast tissue was dense, to what extent did you read the information provided? (%) |  |  |  |
| Not at all | 153 (11.1) | 134 (10.2) | 287 (10.7) |
| Glanced at it | 157 (11.4) | 141 (10.8) | 298 (11.1) |
| Read some parts | 187 (13.5) | 222 (17.0) | 409 (15.2) |
| Read it completely | 867 (62.8) | 769 (58.8) | 1636 (60.8) |
| Missing | 17 (1.2) | 42 (3.2) | 59 (2.2) |
| Do you think the amount of information provided by BreastScreen WA was... (%) | 1211<sup>a</sup> | 1132<sup>a</sup> | 2343<sup>a</sup> |
| Too little | 394 (32.5) | 203 (17.9) | 597 (25.4) |
| Just Right | 570 (47.1) | 650 (57.4) | 1220 (52.1) |
| Too much | 3 (0.2) | 2 (0.2) | 5 (0.2) |
| Don't know | 241 (19.9) | 260 (23.0) | 501 (21.4) |
| Missing | 3 (0.2) | 17 (1.5) | 20 (0.9) |
| In your own words, what did you learn from the information provided?<sup>b</sup> (%) | 1211<sup>a,c</sup> | 1132<sup>a,c</sup> | 2343<sup>a,c</sup> |
| Informed about what to do | 764 (63.1) | 518 (45.8) | 1282 (54.7) |
| Informed about difficulty detecting cancer | 136 (11.2) | 203 (17.9) | 339 (14.5) |
| Informed about breast density | 347 (28.7) | 222 (19.6) | 569 (24.3) |
| Not informed enough in general | 78 (6.4) | 68 (6.0) | 146 (6.2) |
| Not informed enough about what to do | 88 (7.3) | 15 (1.3) | 103 (4.4) |
| Not informed enough about why now do I have dense breasts | 42 (3.5) | 2 (0.2) | 44 (1.9) |
| Not informed enough about how urgent/problematic | 6 (0.5) | 0 (0.0) | 6 (0.3) |
| Worried about being at increased risk of cancer | 1 (0.1) | 0 (0.0) | 1 (0.1) |
| Worried about GP being not well informed | 3 (0.3) | 5 (0.4) | 8 (0.3) |
| Worried in general | 3 (0.3) | 1 (0.1) | 4 (0.2) |
| Can't recall | 34 (2.8) | 78 (6.9) | 112 (4.8) |
| Miscellaneous | 44 (3.6) | 93 (8.2) | 137 (5.9) |
| No comment | 97 (8.0) | 122 (10.8) | 219 (9.3) |

a. Question was not asked for women who answered 'not at all' or who did not respond to the question 'The first time BreastScreen WA told you that your breast tissue was dense, to what extent did you read the information provided?'
b. Categories derived from framework found in Supplementary Table 3
c. Responses can be coded into more than category

Women were invited to comment on what they understood from the information provided by BreastScreen WA and these open-ended responses were analysed (Supplementary Table 3) and summarized in broad categories in Table 3. Around three quarters of women indicated they felt informed, with half of them indicating that they learned what to do in terms of following up with their GP and around 15% indicated they learned that it became more difficult to detect cancer in dense breasts. A small proportion of women indicated that they were not informed enough, particularly about general breast density information or about what to do.

## Discussion

This large survey of women attending a public mammographic screening program is the first to compare knowledge of breast density between women notified of having dense breasts and controls (women not flagged as having dense breasts). We found that over 60% of controls have heard of breast density and would like to know their breast density. Most (>80%) notified women know that dense breast tissue makes it more difficult to see cancer on a mammogram and that sometimes women may require further tests if they have dense breast tissue, compared to just over half of controls (∼55%). Overall (both notified and control groups), women largely do not know that having dense breasts means they are at increased risk of breast cancer, that breast density cannot be determined by feel or touch, or that breast size does not determine a woman’s breast density. Half of notified women thought the information provided to them by BreastScreen WA was sufficient, however those notified for the first time were less satisfied suggesting they may benefit from additional information.

General awareness of breast density in this population (∼60%) is near the top end of estimates from the literature ranging from 39% to 92% (8-15), recognizing that women who have heard of breast density are more likely to complete a survey about breast density. Similar to smaller study (14), we found that a majority of women who do not know their breast density would like to know (72% of controls).

BreastScreen WA’s message appears to be getting through, with 86% of notified women agreeing that dense breast tissue makes it more difficult to see cancer on a mammogram. American estimates of knowledge about the reduced sensitivity of mammography range from 20-66% (8, 12, 13, 16, 17). This variability could be due to wording, reader comprehension, or even the layout of information on the results letter (16). Our population were also knowledgeable about supplementary screening, with 67% of all women agreeing or strongly agreeing that women with dense breasts may require further tests, compared to 34% in a another study (10). When asked if breast density puts women at increased risk of breast cancer, we found that 12% of controls and 25% of notified women responded correctly. This is similar to the findings from a random sample of Virginian women, where only one in eight were aware that density increases breast cancer risk (8). In contrast, a recent study reported that ∼65% of women within a US probability-based sample (oversampled in Connecticut) knew that breast density affects cancer risk (17). BreastScreen WA does not provide information regarding the association between breast density and breast cancer risk in its results letter, however the report includes a link to BreastScreen WA’s dedicated breast density webpage that does provide information about increased risk associated with increased breast density.

This was the first study to ask notified women to what extent they read the information provided by their screening program and what they understood. The analysis of open-ended responses indicated that half of notified women thought that the information provided to them by BreastScreen WA was sufficient and almost 80% of the open-ended comments about what they understood related to feeling more informed. Post-hoc cross-tabulation shows that women with similar levels of understanding reported different levels of satisfaction with information provided. For example, one woman who indicated that the information was “just right” understood “That I had dense breasts and so the mammogram may not be effective and I should see my doctor.” Another woman who indicated that the information was too little responded “I only received a letter that notified me that I have dense breasts and that mammogram is not sufficient for cancer screening because of that. I think they suggested to see my GP and perhaps have further testing.” Finding a balance of information that satisfies the majority and recognises different levels of health literacy and interest is likely to remain challenging.

Study limitations include a response rate of 23.5% however the recruitment target was 6000 completed surveys within a 6 month period and a minimum expected participation rate of 10%. We argue that our response rate was good for an unsolicited email invitation for an online survey. Also, studies have demonstrated that response rates are not directly correlated with validity of results; some studies with low response rates, even as low as 20%, are able to yield more accurate results than studies with response rates of 60% to 70% (18, 19). This survey was population-based, nested within a public breast cancer screening program in Australia so its generalisability is dependent on screening programs and healthcare systems of other populations.

This study demonstrates relatively high levels of knowledge within a population-based screening program, particularly with respect to reduced sensitivity of mammography in women with dense breasts. It also highlights some of the challenges when relaying health information to large populations; it is difficult to provide sufficient information to everyone, particularly when key information regarding what women should do is unclear. However, this study identified potential areas for improvement particularly, more information for women notified that they have dense breasts for the first time.

## Data Availability

The datasets generated and analysed during the current study are not publically available but can be made available upon request (and pending
approval) from BreastScreen Western Australia.

## Conflict of Interest

The authors declare no conflicts of interest.

## Notes

### Competing Interest Statement

The authors have declared no competing interest.

### Funding Statement

This work was supported by Cancer Australia (APP1085750) and the Commonwealth of Australia Public Health and Chronic Disease Grant Program. JS is a National Breast Cancer Foundation Research Fellow.

